# Feasibility of an adapted participatory group programme for caregivers of children with complex neurodisability in the United Kingdom: Results from the Encompass-2 study

**DOI:** 10.64898/2026.02.11.26346106

**Authors:** Kirsten Prest, Kirsten Barnicot, Catherine Hurt, Cally J Tann, Michelle Heys, Angela Harden

**Author notes:** MH and AH should be considered joint last authors.

## Abstract

**Introduction:** “Encompass” is a participatory group-based intervention originating from low- and middle-income countries, co-developed with parents and professionals to enhance the wellbeing, health literacy and empowerment of caregivers of young children with complex neurodisability. We aimed to assess feasibility and acceptability of a) intervention delivery in two socially deprived United Kingdom (UK) urban areas and b) evaluation methods including data collection on programme outcomes and costs.

**Methods:** We conducted a mixed-methods pilot and feasibility study with caregivers of children under five years with complex neurodisability. Feasibility and acceptability of intervention delivery were assessed based on recruitment rates, group attendance, fidelity checklists and qualitative interviews with caregivers and facilitators. Feasibility and acceptability of evaluation methods were explored through follow-up rates, questionnaire completeness, and caregiver feedback on outcome measures. Data relating to implementation at organisational and system levels were explored through interviews with facilitators and key partners. Results were compared to predefined traffic light criteria (green, amber, red) to determine whether a larger scale evaluation was warranted.

**Results:** Eight caregivers participated in the programme. Fidelity of delivery and follow-up questionnaire completion met green criteria, while recruitment and attendance met amber criteria, indicating that minor adaptations are required before scaling up. Qualitative findings demonstrated high acceptability of the programme among caregivers and facilitators, particularly valuing the co-facilitation model, participatory approach, and peer support. Flexible delivery, including online participation and communication support, enhanced accessibility for families with diverse needs. Capturing programme delivery costs was feasible and provided preliminary estimates to inform future economic evaluation.

**Conclusions:** Our findings provide proof of principle that “Encompass” can feasibly and acceptably be delivered and evaluated with caregivers of children with complex neurodisability in an ethnically diverse UK community health setting. The findings support progression to a larger-scale evaluation, with refinements to recruitment strategies and delivery logistics.

**Patient or Public Contribution:** Caregivers with lived experience were central to developing the “Encompass” programme and this study. Four local mothers of children with complex neurodisability contributed to planning, recruitment, and sense-checking the findings.

## Introduction

The task of caring for a child with a complex neurodisability places unique and considerable demands on their caregivers, frequently resulting in caregivers having unmet physical, psychological, information and social needs (1–5). Many caregiver needs reflect systemic barriers to caring for a child in a society with health and education systems that pose fundamental barriers for those with impairments (6). For example, lack of time for themselves, managing complex, fragmented health systems and numerous appointments, financial stress, and the social impact on dynamics with family and friends (7–14). Caregivers also face challenges related to managing their grief and acceptance (7–9), and the physical effort required to care for their child (10,11). In this paper, we use the term “caregiver” to refer to parent carers, relatives or anyone who provides primary care to the child. We use the term “complex neurodisability” to describe children with motor disorders that have a neurological cause. For example, cerebral palsy (CP), an umbrella term used to describe disorders caused by an injury to the developing brain resulting in difficulties with movement and posture along with participation and activity limitations (15). CP affects 1 in 400 children in the United Kingdom (UK) (16).

Family-centred care, which involves working collaboratively with caregivers to support the individual needs of families (17), is the gold standard in healthcare for all children including those with complex neurodisability. However, it is often not implemented effectively, and many families require further support, including information about their child’s condition and health services available, and how to connect with other parents (18,19). Along with the provision of holistic, family-centred care, peer support is an important protective factor for the health and wellbeing of these caregivers (20,21) with group programmes having been shown to improve their skills, knowledge, wellbeing and empowerment (22). UK group-based programmes such as “E-PAtS” (Early Positive Approaches to Support) and “Healthy Parent Carers” have shown to improve wellbeing, peer support, and service navigation for caregivers of children with disabilities (23–25). However, they are not tailored to the specific needs of caregivers of children with complex neurodisability. A gap therefore remains in early-years support addressing physical care demands, condition-specific information, and specialist pathways. “Encompass” was developed to address this gap as a structured, co-delivered programme for families of young children with complex neurodisability.

“Encompass” is based on the “Baby Ubuntu” programme which is widely implemented across low- and middle-income countries (26). It is an example of a ‘decolonised healthcare innovation’ as it acknowledges that low-cost, community-based solutions developed in resource-constrained settings may be relevant for high-income countries like the UK. “Encompass” was developed based on a qualitative study (Encompass-1) exploring caregiver needs in the local communities where this study was based (27). “Encompass” is a ten-module participatory group programme for caregivers of children with complex neurodisability under 5 years of age. It is co-facilitated by a parent with lived experience along with a healthcare professional (26,28).

This paper reports on initial feasibility and pilot testing of the “Encompass” programme for caregivers of children with complex neurodisability under 5 years in two urban areas with high levels of social deprivation and ethnic diversity in the UK (referred to as the Encompass-2 study). The study aimed to a) assess the feasibility and acceptability of delivering the programme (i.e. recruitment, retention, fidelity, acceptability) and b) pilot evaluation methods to inform a future larger-scale evaluation of “Encompass” (i.e. data collection methods for assessing programme outcomes and costs, completion of baseline and follow-up). As part of the study, data on the perceived impact of the programme were also collected but these are reported in a separate paper.

## Materials and Methods

### Design

This is a mixed methods single group pre- post pilot and feasibility study that aimed to determine the feasibility and acceptability of delivering and evaluating the “Encompass” programme to caregivers of children under five years with complex neurodisability. Detailed methods can be found in the study protocol (28) and the study registration on clinical trials.gov (ID: NCT06310681).

### Setting

The study is set in two ethnically, culturally and linguistically diverse London boroughs in the UK. A borough refers to a local government district, typically home to several hundred thousand residents (29). The populations in these boroughs face challenges with language barriers, health inequalities and approximately one in two children are considered to be living in poverty (30–32). These boroughs have higher rates of low health literacy (58–67%) than the UK average (41%), creating challenges for navigating health information and services (33) and contributing to poorer health outcomes (34).

### Intervention

The “Encompass” caregiver programme follows ten modules covering a range of topics (see figure 1). “Encompass” utilises the ‘expert parent’ facilitator model (35) and provides an opportunity for caregivers to learn from each other’s experiences, using participatory approaches and principles of adult learning (36). Children were invited to the groups so that caregiver skills could be practised. A full description of the programme, framed within the Template for Intervention Description and Replication (TIDieR) checklist, can be found in Prest et al. (2025) (26).

**Figure 1:**
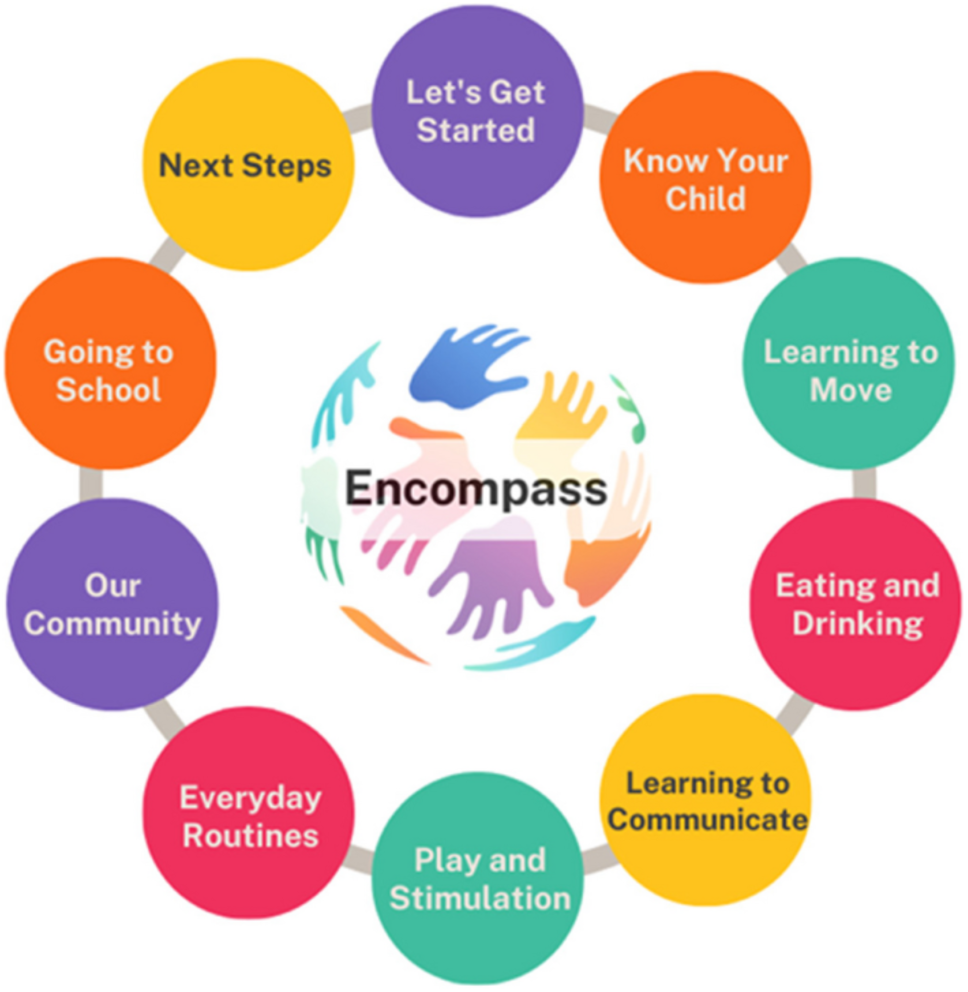
The “Encompass” modules reproduced from Prest et al. (2025) (26)

### Involvement of those with lived experience

The involvement of those with lived experience has been an essential component throughout the development of the “Encompass” programme and has remained a core aspect of this pilot and feasibility study. Four local mothers of children with complex neurodisability were involved in the planning stages of this study (reviewing participant-facing documents), the implementation (advising on recruitment and advertising the study), and analysis (sense-checking findings).

### Participants and recruitment

The study population included the following groups: (1) Caregivers of children with complex neurodisability under 5 years who resided in the study area, (2) the children of the above caregivers, (3) the facilitators responsible for the delivery of the programme (healthcare professionals and expert parents) and (4) key partners within the NHS and other areas involved in the delivery or commissioning of health and care services to the local population.

Caregivers of children with complex neurodisability who met the inclusion criteria (see table 1) were invited to participate in the study during appointments with trusted clinicians or via telephone calls made by those involved in early years therapy teams (e.g. occupational therapists, physiotherapists or speech and language therapists). They were given a participant information sheet and the contact details of the researcher during these interactions. Participants expressed their interest via a QR code or by emailing the researcher directly. Informed consent was obtained after they had sufficient time for any questions to be addressed.

**Table 1:** Inclusion and exclusion criteria.

|  | Inclusion | Exclusion |
| --- | --- | --- |
| Caregivers and children | Caregivers who:<br>Cared for a child (<5 years at the time of enrolment) with a complex neurodisability<br>Had received a diagnosis for their child, which had been disclosed to them, even if this was a diagnosis such as SWAN (Syndrome Without A Name)<br>Resided in the two study boroughs<br>≥18 years of age. | Caregivers who cared for a child with:<br>A developmental disability where there were no functional physical impairments as part of their complex needs. For example, children diagnosed with Autism Spectrum Disorder, Attention Deficit Hyperactivity Disorder, intellectual impairments were excluded unless they had a functional physical impairment with a neurological cause too.<br>A progressive neurological condition such as Duchenne’s Muscular Dystrophy as the family’s needs would be different from those whose child had a life-long condition but was not progressive.<br>A structural physical impairment not caused by a neurological event or neurological difficulties. For example, children born with a limb difference or a child with hearing loss.<br>Did not have capacity to consent.<br>There were no exclusions based on language, as interpreting/translating services were offered. |
| Facilitators | (1) Healthcare professionals:<br>Therapists or healthcare professionals who were confident in working with children with complex neurodisability and who are open to working alongside and learning with families about their children.<br>(2) Expert parents:<br>Parent or caregivers of a child with complex neurodisability<br>Previous experience in facilitating was not essential, but they had to be open to working alongside a healthcare professional and show | Inability to read and speak English<br>Inability to commit to a 6-month period of work to the best of their knowledge |
|  | an understanding of participatory approaches. |  |
| Key NHS Partners | Key partners in the local health and care system (such as clinical managers or commissioners)<br>Community workers in the across health, social care and voluntary sector |  |

Healthcare professional facilitators were recruited from the two community child health services serving the study areas. Expert parents were identified through the research parent partners and through local clinicians.

Key partners were approached via local collaborators to be interviewed to gain their perspectives on the feasibility of implementing a programme like “Encompass” in their local settings.

### Data Collection

Data collection methods and corresponding analyses were organised around five feasibility objectives, summarised in table 2.

**Table 2:** Data collection and analysis methods for feasibility objectives.

| **Feasibility objective** | **Data collection** | **Data analysis** |
| --- | --- | --- |
| Feasibility of recruitment, retention, and fidelity of the intervention | Recruitment and retention logs, attendance registers, fidelity checklists completed by KP (and by a second reviewer for 20% of sessions). | Recruitment and retention rates calculated descriptively, fidelity checklist items scored and converted to percentages, both assessed against pre-specified traffic-light criteria (green, amber, red) for progression. |
| Acceptability of the intervention from the perspectives of facilitators and caregivers | Semi-structured interviews with caregiver participants and facilitators, satisfaction questionnaire using Likert scales, feedback on facilitator training and manual. | Descriptive qualitative analysis (inductive coding to identify categories relating to acceptability), quantitative questionnaire data summarised descriptively. |
| Feasibility and acceptability of the intervention at a system and organisational level | Semi-structured interviews with facilitators and key partners using topic guides informed by the Context Compass framework. | Deductive coding guided by the Consolidated Framework for Implementation Research (CFIR), focusing on inner and outer setting domains. |
| Questionnaire response rate, completeness, and acceptability of outcome measures | Follow-up response rates, caregiver interview feedback on outcome measures. | Quantitative completion rates summarised descriptively, qualitative feedback presented narratively to describe perceived acceptability. |
| Feasibility of assessing the cost of the programme | Programme delivery costs captured using the Childhood Cost Calculator, | Costs summarised descriptively by category (labour/personnel, |
|  | categorised by investment and recurrent costs. | materials/supplies/equipment, facilities). |

Quantitative data collection methods included recruitment logs, group attendance, a fidelity checklist adapted from the one used by the “Baby Ubuntu” team (supplementary file 1), a Likert scale questionnaire measuring satisfaction, response rates for outcome measure questionnaires, and recording the cost of the programme using the Childhood Cost Calculator (37).

Qualitative data were collected through semi-structured interviews with caregiver participants, facilitators, and key partners. Separate topic guides were developed for each participant group (supplementary file 2). Interviews were conducted by KP, an occupational therapist trained in qualitative interviewing, primarily online or in caregiver participants’ homes, and were audio-recorded, transcribed, and pseudonymised. Interviews lasted between 40 and 70 minutes.

### Data analysis

Quantitative data relating to feasibility of recruitment, retention rates and fidelity were assessed against predefined traffic light criteria for progression to a larger scale evaluation. Green ratings indicated that thresholds were met and progression to a larger evaluation would be feasible, amber suggested that adaptations may be required, and red indicated that the criteria were not met. Thresholds for each domain were pre-specified in the study protocol (28).

Recruitment rates were calculated based on how many caregivers consented to the study of those who were approached and were eligible to participate. Retention rates were calculated based on how many groups (out of 10) each participant attended. Items on the fidelity checklist included observations relating to the logistics of the group, the preparation and the facilitation skills (supplementary file 1). The fidelity checklist was completed for each group by KP, with a second reviewer for 20% of groups. Checklists were completed independently, discrepancies resolved by consensus, and group scores totalled and converted to percentages. Quantitative data relating to response rates and completeness of outcome measures were presented descriptively. The costs of delivering the programme were divided into investment and recurrent costs, as well as resource types (37).

The qualitative interview transcripts were coded and analysed descriptively by KP, both inductively and deductively, with results discussed with the wider team to improve reflexivity and consistency. Data were managed using NVivo software (38). Inductive descriptive qualitative analysis involved coding interview transcripts from caregivers and facilitators to identify and interpret key categories related to feasibility and acceptability (39). Deductive analysis was guided by the Consolidated Framework for Implementation Research (CFIR), with transcripts from facilitators and key partners coded to inner and outer setting domains to examine feasibility at system and organisational levels (40). Interview guides were informed by the Context Compass framework, which is partly based on CFIR (41), however, CFIR was used for analysis because Context Compass was still under development. Qualitative data about the acceptability of outcome measures were presented descriptively.

### Ethics

Ethical approval was obtained from the Health Research Authority (ref. 23/EM/0213). Procedures for informed consent were followed to ensure all participants (caregivers, facilitators and key partners) understood what their participation would involve before agreeing to be a part of the study. Measures were in place to ensure confidentiality was maintained (along with the limits of confidentiality), safeguarding policies were adhered to, and facilitators were trained to support vulnerable caregivers who may be displaying signs of emotional distress. All personal data were collected and stored in accordance with the Data Protection Act 2018 and General Data Protection Regulation.

## Results

The results are presented in five sections corresponding to the feasibility objectives that examined the feasibility and acceptability of both delivering and evaluating the “Encompass” intervention. These include (1) the feasibility of recruitment, retention, and intervention fidelity, (2) the acceptability of the intervention from the perspectives of caregivers and facilitators, (3) the feasibility and acceptability of the intervention at system and organisational levels, (4) questionnaire response rate, completeness, and the acceptability of outcome measures, and (5) the feasibility of capturing programme costs. Illustrative quotes are provided throughout the results with C= caregiver participant, F= facilitator (either parent or professional) and KP= key partner.

### Feasibility of recruitment, retention, and intervention fidelity

#### Recruitment

Out of 35 caregivers who were eligible and approached for the study, eight provided their consent to participate. Figure 2 shows the flow of participants through the study and sociodemographic information for children and caregivers is shown in table 3.

**Figure 2:**
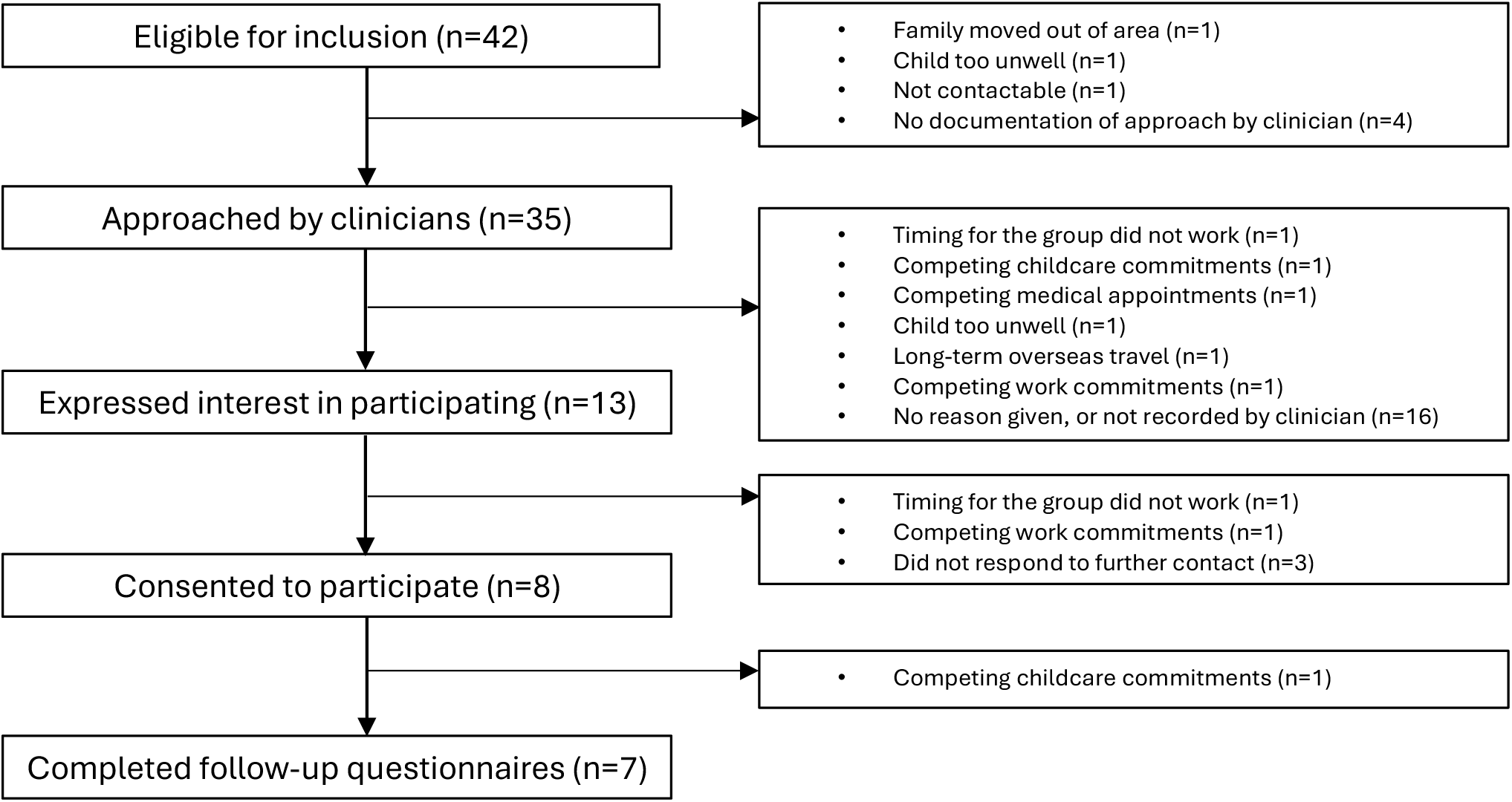
Flow of caregiver participants through the study

**Table 3:**
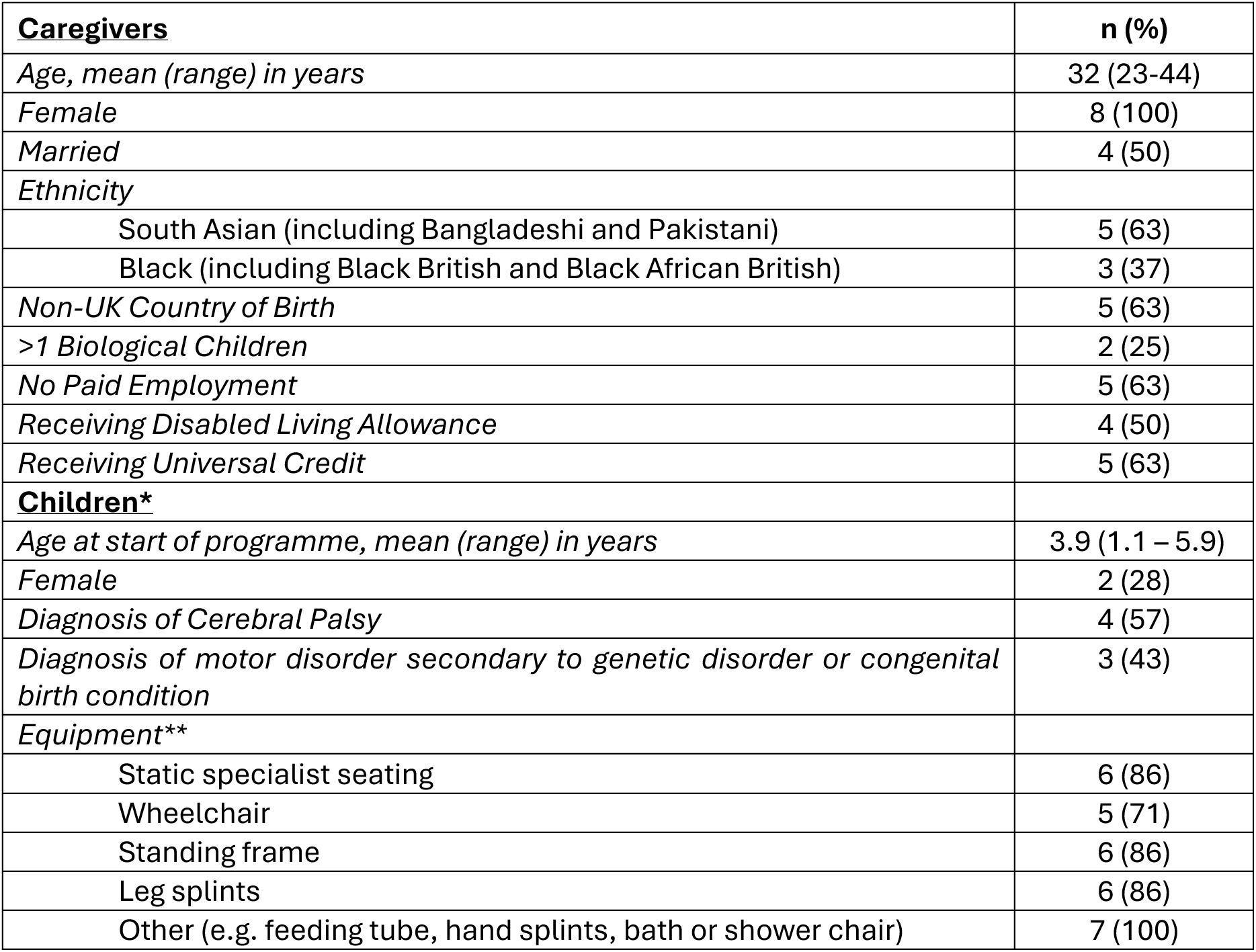
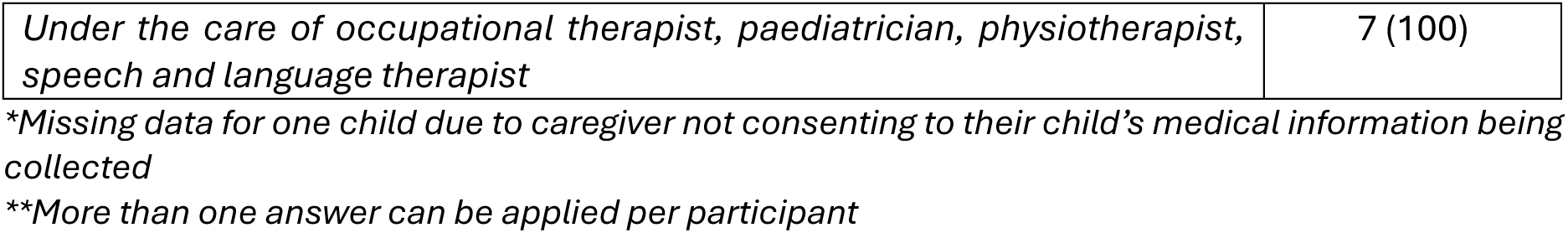
Demographic information of caregivers and children (N=8)

| **Caregivers** | **n (%)** |
| --- | --- |
| Age, mean (range) in years | 32 (23-44) |
| Female | 8 (100) |
| Married | 4 (50) |
| Ethnicity |  |
| South Asian (including Bangladeshi and Pakistani) | 5 (63) |
| Black (including Black British and Black African British) | 3 (37) |
| Non-UK Country of Birth | 5 (63) |
| >1 Biological Children | 2 (25) |
| No Paid Employment | 5 (63) |
| Receiving Disabled Living Allowance | 4 (50) |
| Receiving Universal Credit | 5 (63) |
| **Children\*** |  |
| Age at start of programme, mean (range) in years | 3.9 (1.1 – 5.9) |
| Female | 2 (28) |
| Diagnosis of Cerebral Palsy | 4 (57) |
| Diagnosis of motor disorder secondary to genetic disorder or congenital birth condition | 3 (43) |
| Equipment** |  |
| Static specialist seating | 6 (86) |
| Wheelchair | 5 (71) |
| Standing frame | 6 (86) |
| Leg splints | 6 (86) |
| Other (e.g. feeding tube, hand splints, bath or shower chair) | 7 (100) |
| *Under the care of occupational therapist, paediatrician, physiotherapist, speech and language therapist* | 7 (100) |

*\*Missing data for one child due to caregiver not consenting to their child's medical information being collected*
*\*\*More than one answer can be applied per participant*

Eight of 35 (23%) eligible caregivers consented to participate in the study, meeting amber light criteria (table 4). The main reasons reported for declining participation were clashing commitments (e.g. attending other groups or work schedules) and concerns about the child being too unwell to attend, however reasons for not participating were not consistently reported by local clinicians. One of the caregiver participants in the study described how some families may not be emotionally ready for a group like “Encompass” and although this reason was never given explicitly, it may be an indication as to why caregivers chose not to participate.

> *“I think that’s why [some parents] don’t want to come. Our emotions are so high and intense that we are literally moving in fight mode, even when we’re sleeping… so to have our emotions constantly bottled up, the fear of coming to a group and opening that means you’re gonna be vulnerable.” (C6)*

**Table 4:** Traffic light criteria to progress to a larger scale evaluation.

|  | **Traffic light criteria** | **Results** | **Traffic light** | **Interpretation** |
| --- | --- | --- | --- | --- |
| **Recruitment** | Green: 35% consent<br>Amber: 15-34%<br>Red: <15% | 8 out of 35 (23%) caregivers who were approached consented to participate | Amber | Amendments required before progression |
| **Group attendance** | Green: ≥80% attend<br>≥6 sessions<br>Amber: 30-79%<br>Red: <30% | 5 out of 8 (63%) caregivers attended ≥6 sessions | Amber | Amendments required before progression |
| **Follow-up response rate** | Green: ≥70% response rate<br>Amber: 50–69%<br>Red: <50% | 78% (94/120*) response rate | Green | Criteria for progression met |
| **Fidelity** | Green: ≥70% score<br>Amber: 50–69%<br>Red: <50% | Mean score of 87% on the fidelity checklist | Green | Criteria for progression met |

\*Response to 5 outcome measures over 3 timepoints (pre, post and follow-up) for all 8 participants.

Recruitment was most effective when the study was discussed with the potential participant by a trusted clinician. Using a QR code to register interest was more effective than asking caregivers to email the researcher directly.

In the qualitative interviews, participants and facilitators gave recommendations about how to reach more families to tell them about the “Encompass” groups. One recommendation was using word of mouth of caregivers who had attended the group, as they might be more likely to listen to other caregivers with similar lived experience. Others suggested advertising the groups through local community workers and schools.

> *“When it comes from therapists, parents do feel like, ‘oh, it’s another appointment, something that I have to go to’. But when it comes from parents who’ve been to that workshop and they benefited, they have met other parents or connected with other parents, they’d be like ‘I went there, and it was helpful’” (F2 - parent)*

#### Retention

The study began with two groups running separately. However, after five modules had been delivered, the groups were combined due to small numbers of participants. After the first group was held, one participant dropped out of the study due to having another baby. They have been included in the analysis to reflect real world scenarios, even though they only attended one group. This means 5 out of 8 caregivers (63%) attended six or more out of the ten modules (amber light criteria, table 4). Reasons given for non-attendance were competing medical appointments for the children, a child being unwell, or work commitments.

#### Fidelity

Groups were run with high fidelity according to the fidelity checklists completed. There was a mean score of 87% on all fidelity checklists completed (green light criteria, table 4). Items that facilitators consistently scored high on included the room set up, facilitators asking participants to share their own experiences, using verbal and non-verbal communication and body language, and being prepared and knowledgeable about the content. Items that at times scored lower included facilitators summarising discussions, asking the group if they had any questions after each activity, and following every component in the manual.

### Acceptability of the programme from the perspectives of caregivers and facilitators

This section presents findings related to the acceptability of the “Encompass” programme, drawing on caregiver and facilitator perspectives. Based on a self-report satisfaction scale, caregivers who attended the programme were highly satisfied with its content, organisation, and facilitation (table 5). Three descriptive categories were developed inductively from the qualitative analysis: (1) *Facilitation grounded in lived experience*, (2) *Relevant, practical, and engaging content*, and (3) *Flexibility and inclusion enhance participation*.

**Table 5:** Satisfaction with the content, organisation and facilitators of “Encompass” from the caregivers’ perspective.

| Feedback domain | Score* (n=6) |
| --- | --- |
| Satisfaction with the content of the programme | 4.8 |
| Satisfaction with the organisation of the programme | 4.8 |
| Satisfaction with the facilitators of the programme | 5 |

\*1= not at all satisfied, 5 = extremely satisfied

#### Facilitation grounded in lived experience

Facilitators and caregivers described the programme’s facilitation as a key element underpinning its acceptability. The co-facilitation model, pairing a healthcare professional with an expert parent, was viewed as unique and valued by both groups. Facilitators also reflected that the training supported their confidence and ability to deliver the programme.

Healthcare professionals described working alongside expert parents as “inspiring,” while expert parents valued the partnership and mutual respect.

> *“It was really nice working with a parent. They were so passionate about it… I felt that was really empowering for me and for them, and for the group. It made it more relaxed.” (F3 - professional)*

> *“Obviously they didn’t have children with a disability, but they’ve worked with children with disabilities for a long time in their careers…I really enjoyed working alongside them.” (F1 - parent)*

Caregivers also recognised this dynamic as something that made the programme distinctive and credible. They valued hearing from someone with lived experience who could offer empathy and realistic advice alongside professional guidance.

> *“To then have a parent stand there and talk, being in their power, but then also coming with the realistic side of their life, that’s what we want. We want it to be authentic.” (C6)*

Caregivers appreciated the facilitation style and welcoming nature of the group, which allowed them to feel safe to share their experiences.

> *“The facilitators were really understanding. I felt like they actually listened to us and didn’t just talk at us. It made it easier to open up.” (C1)*

The facilitator training lasted three days and activities were based on the “Baby Ubuntu” facilitator training manual. Facilitators described the training as interactive and collaborative, mirroring the participatory ethos of “Encompass”.

> *“And we did the whole thing as a group, [including] that ice breaker, as if we were the learners. So that was really realistic, you could see it from their perspective.” (F4 - professional)*

Although some felt that three days was not enough to cover all ten modules, most felt it provided a strong foundation and boosted their confidence.

> *“Even though we didn’t cover all the modules, I still think it gave us the confidence to carry on… because we knew how each module worked.” (F1 - parent)*

Due to the pragmatic decision to combine the groups halfway through the programme, both expert parents stayed involved while the healthcare professionals alternated sessions. This meant that there were at times three facilitators leading the group. All facilitators agreed that this was not ideal, however it was seen as a positive for the caregiver participants as they were able to grow their support network and meet more people.

> *“I didn’t feel like having all of us there worked personally. I think it was much better one therapist and one parent. I think it was like too many chefs in the kitchen.” (F4 - professional)*

#### Relevant, practical and engaging content

Facilitators generally viewed the “Encompass” manual as a useful tool that helped them prepare for sessions. Most found it clearly laid out and easy to navigate, though some felt it was overly detailed or included terminology that could be difficult for non-professionals.

> *“With the [manual], I found that some words I struggled to understand. Because I’m coming from a parent perspective, not a professional who knows big fancy words.” (F2 – parent)*

Caregivers particularly valued the balance between structured content and open discussion, noting that the flexibility to share experiences made sessions more meaningful.

> *“I quite enjoyed having the structure of the topic, but then also just going a little bit off on a tangent when people were sharing experiences… I thought that was really good because people voiced concerns that I had as well.” (C5)*

Ice breaker activities were highlighted by both facilitators and caregivers as particularly effective in building rapport and encouraging open discussion, especially when sessions addressed emotionally sensitive topics.

> *“I think the ice breakers are definitely important… they were the parts that people usually opened up about their emotions.” (F3 - professional)*

> *“The string ice breaker activity, that was great in terms of emotional expression… it really demonstrated my feelings as a parent, what my perspective is, how I link that to my child.” (C3)*

Caregivers consistently described the session content as directly applicable to their daily lives. Topics such as Education, Health and Care Plans (EHCPs), learning to communicate, access to community support, and practical demonstrations on eating, drinking, and positioning were all described as useful and relevant.

> *“I found the sessions really practical, things I could actually use at home.” (C7)*

> *“I remember the day we discussed about difficulties with transport, and feeding as well… that helped as well.” (C8)*

#### Flexibility and inclusion enhance participation

Participants described how the flexible delivery of the “Encompass” programme supported engagement and attendance. The timing of the group sessions was also viewed positively.

> *“That slot during the day was like a perfect time when you wake up and you do all your routines and then you go.” (C2)*

Participants could join online when unable to attend in person, which supported continued participation around work and caregiving demands. While they valued this flexibility, they reported reduced social connection compared with in-person sessions and noted that online access may be more challenging for those less familiar with video technology.

> *“The only downside to not being in the room is the extra conversations that happen during breaks… there was one session where you forgot to mute the computer, and it was really interesting to hear the background conversation… it’s those organic discussions that you miss when you’re online.” (C5)*

A caregiver participant required a sign-language interpreter, and this was the first time they had been able to join a group programme. They had worries before about attending the group, however these were appeased after attending the first session. Not only was the interpreting helpful in promoting participation, but the actual activities were visual and practical which were useful for anyone whose first language was not English.

> *“I think the [activities in the group were] useful for me because I could visually see the demonstrations in terms of physical movement. There was a lot also in terms of body language movement. For me as a deaf person, I need to get like a clear picture of how things actually work and that’s really important for me.” (C3)*

The presence of sign language interpreters not only accommodated the caregiver participant who required this, but also prompted reflection and inspiration among other caregivers whose children might similarly benefit from such support.

> *“When you had the interpreters at the group, like that was really, really good. And the way I was just looking at the lady thinking ‘Oh my God, this is amazing’”. (C1)*

### Feasibility and acceptability of the intervention at a system and organisational level

Three key partners (along with the “Encompass” facilitators) were interviewed to explore the feasibility of implementing the “Encompass” programme within their local contexts. They included (1) a clinical manager of community children’s therapy services, (2) a programme lead responsible for strategy and commissioning for babies, children and young people within the regional health system, and (3) a lead community connector working in the voluntary sector to provide support to the local residents (including running support groups for parents). Findings from these interviews are presented within the Consolidated Framework for Implementation Research (CFIR).

#### Outer Setting Domain

The outer setting explores the broader contextual and system factors influencing the feasibility and acceptability of implementing “Encompass”.

##### Partnerships and connections

Partnerships and connections across the NHS, social care, and education were viewed as a strength for potential implementation. Key partners mentioned the importance of strengthening collaboration with various partners to reach diverse families and build trust.

> *“Residents may not trust the government or even the NHS, but they do trust their local clinicians and community leaders, faith leaders, [and] voluntary and community sector organisations that they have regular contact with.” (KP2)*

However, there were differing perspectives about the most appropriate setting to host “Encompass”. Some facilitators felt that children’s centres would be a suitable environment while others argued that the programme’s clinical elements required closer integration with children’s therapy teams to ensure governance and supervision.

> *“You wouldn’t want to hand it over to children’s centres… you’re discussing clinical things, swallowing, positioning… it’s more in line with therapeutic [aims], especially when they’re so medically complex and so sensitive.” (F4 - professional)*

> *“I think maybe going into children’s centres and working with the staff there perhaps could work.” (F3 - professional)*

These perspectives suggest that partnerships and connections could act as both facilitators and potential barriers. While community links may increase accessibility and reach, uncertainty around ownership of the programme need to be addressed.

##### Commissioning priorities and financing

Key partners described national and local policy priorities, particularly around early intervention and equity in children’s services, as supportive of the objectives of “Encompass”.

> *“Children have been underfunded in the NHS compared to adults… it’s something that I’m very passionate about, ensuring that we get the best offer across [the local area]” (KP2)*

However, NHS financing and commissioning processes may act as barriers to the feasibility of implementing a sustainable longer-term intervention.

> *“Our trust has got financial difficulties… most of the year you go around having like no budget and then suddenly you get a panic email which says, ‘We’ve got this really great opportunity, but you need to do something by tomorrow night.’” (KP1)*

##### Local conditions and attitudes

The local attitudes and conditions could present barriers to the implementation of “Encompass” and may hinder caregivers’ participation in the groups.

> *“I think we’ve seen a lot of people struggling with energy bills and with rent increases. A lot of people struggling with making ends meet…and we see social isolation of residents who come in.” (KP3)*

> *“We also have a significant movement of population… you’ve got a very transient population.” (KP2)*

Another barrier in terms of local attitudes is that people in the population may take time to build trust with the wider NHS and governmental organisations. It is recommended that trusted community leaders be engaged when introducing an innovation. This aligns with caregiver participants’ suggestions to rely on word of mouth within the community to raise awareness of “Encompass”.

> *“It’s really about… understanding how different groups receive their information and advice and who they trust in delivering that information.” (KP2)*

#### Inner Setting Domain

The inner setting domain explores factors at an organisational level relating to the feasibility and acceptability of delivering “Encompass”. Key partners therefore commented on factors relevant to their local NHS-funded children’s therapy services.

##### Constructs inherent to the inner setting

Stable staffing, strong collaboration, and effective team communication were seen as key facilitators for implementation.

> *“I’m really proud of the fact that we are collaborative… there’s leadership at the top, but it doesn’t feel like it’s a dictatorship… people are given space and time to get on board with things or to shape and change things.” (KP1)*

The collaborative, flexible, innovative culture within the team described above was seen as a facilitator to implementing new innovations, but also as a barrier because projects can at times be overly ambitious. Both teams shared strong values around caregiver support (recipient-centredness), emphasising the importance of empowering caregivers over time.

> *“The NHS can’t always be holding [the caregiver’s] hand… we want them to be self-advocating… we need to give them the power or permission… to get support for themselves.” (KP1)*

##### Constructs specific to the delivery of “Encompass”

The most frequently mentioned constructs within the inner setting relating to delivery were compatibility, mission alignment, and relative priority. All participants described how “Encompass” aligned with the children’s community therapy teams’ values and ethos, also noting its fit with the coaching approach already embedded in team priorities.

> *“The [community children’s therapy] team are the people I’d go to for day-to-day things… they combine all three therapies, and “Encompass” feels like an overarching fit, even though it covers more than just the therapies.” (C2)*

> *“It aligns a lot with what we’re already doing and with our values and ethos.” (KP1)*

Facilitators noted that “Encompass” could help prepare caregivers for therapy appointments and improve understanding of different professional roles.

> *“Encompass would make things easier for professionals too… parents would come in with more knowledge, rather than spending two or three appointments just explaining what’s going to happen.” (F1 - parent)*

While teams were positive about the fit, managers emphasised the need to secure buy-in and protect time, so it was not perceived as an add-on.

> *“We need to really get people on board with it, otherwise they see it as maybe something else that they have to do… really getting them to see the value.” (KP1)*

Competing priorities and staffing realities meant introducing “Encompass” may require trade-offs and considering the experience and skills of the staff would be required.

### Questionnaire response rate, completeness, and acceptability of outcome measures

#### Questionnaire response rates and completeness

The questionnaire return rate across three timepoints was 78% (amber light criteria, table 4 with details in table 6). Missing data reflected real-world barriers, including non-attendance, caregiving demands, postal loss, and participant withdrawal. When participants returned their questionnaires, either in-person or by email, they were completed with very little missing information.

**Table 6:**
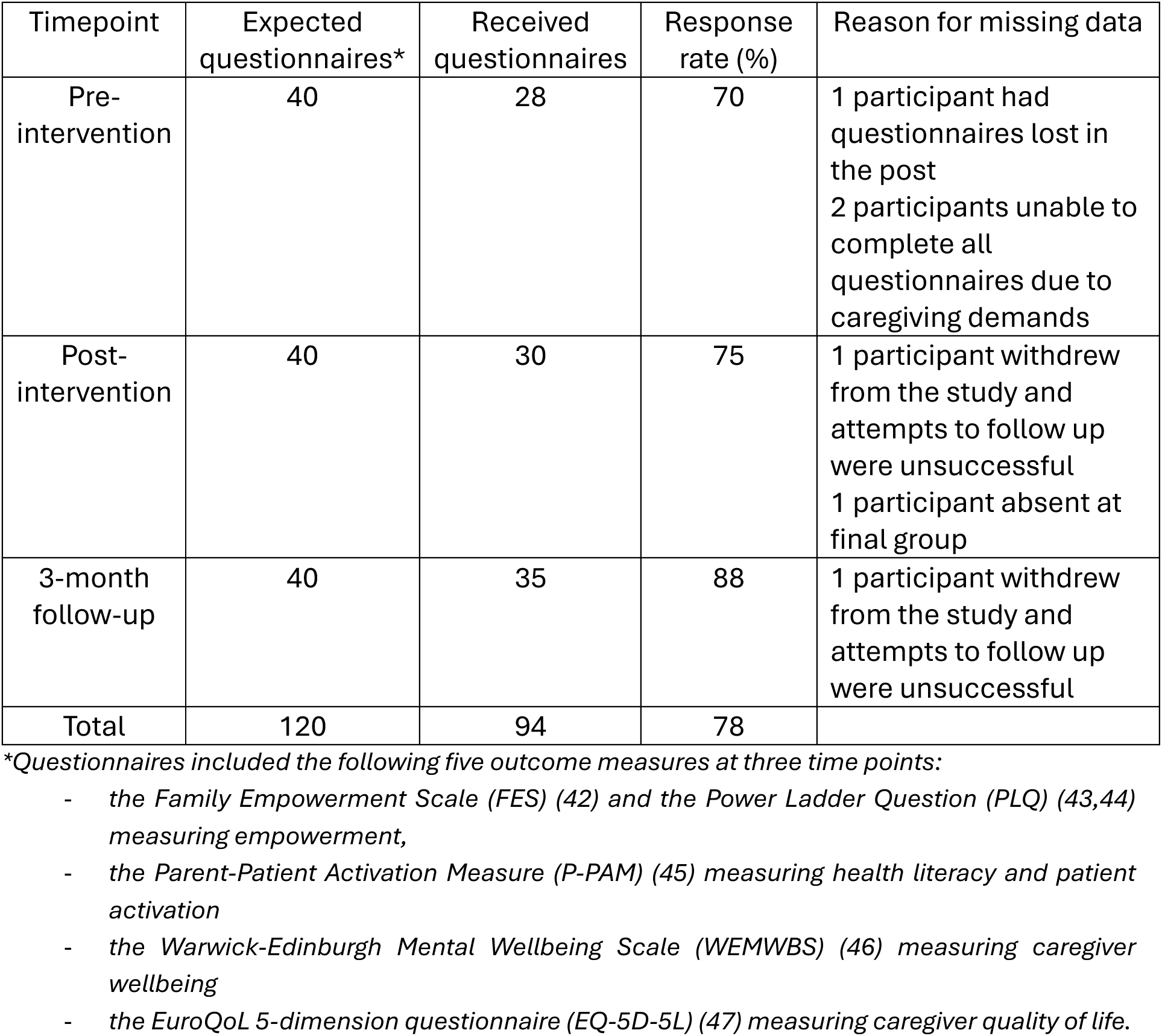
Questionnaire response rates and completeness (N=8)

| Timepoint | Expected questionnaires* | Received questionnaires | Response rate (%) | Reason for missing data |
| --- | --- | --- | --- | --- |
| Pre-intervention | 40 | 28 | 70 | 1 participant had questionnaires lost in the post<br>2 participants unable to complete all questionnaires due to caregiving demands |
| Post-intervention | 40 | 30 | 75 | 1 participant withdrew from the study and attempts to follow up were unsuccessful<br>1 participant absent at final group |
| 3-month follow-up | 40 | 35 | 88 | 1 participant withdrew from the study and attempts to follow up were unsuccessful |
| Total | 120 | 94 | 78 |  |

\*Questionnaires included the following five outcome measures at three time points:
- the Family Empowerment Scale (FES) (42) and the Power Ladder Question (PLQ) (43,44) measuring empowerment, - the Parent-Patient Activation Measure (P-PAM) (45) measuring health literacy and patient activation - the Warwick-Edinburgh Mental Wellbeing Scale (WEMWBS) (46) measuring caregiver wellbeing - the EuroQoL 5-dimension questionnaire (EQ-5D-5L) (47) measuring caregiver quality of life.

#### Acceptability of outcome measures

All caregivers who participated in an interview were asked about the acceptability of the outcome measures. Many caregivers had English as their second language and therefore required some support in filling out the questionnaires in-person and explaining some of the concepts. They described how it was easy to fill them in with support.

> *“Oh, yes I found [filling them in] okay. It was a bit difficult and at least you were there to help me. Yes, because I speak French and there’s a lot of words” (C8)*

> *“They were quite clear to understand. And also the language being clarified as well [helped]. So I think I’ve understood it all very clearly.” (C3)*

Most caregiver participants reported that the Parent-Patient Activation Measure (P-PAM) was the easiest questionnaire to complete. Some participants found the Power Ladder Question (PLQ) difficult to understand, whereas others found it a straightforward and appropriate way to capture their feelings of empowerment. When completing the Family Empowerment Scale (FES) ‘about your involvement in the community’ sub-section, most participants required support in understanding the term ‘legislators’ and how this would relate to their own context and community.

One caregiver reflected on the challenges of completing the pre-intervention questionnaires, explaining that some of the concepts were unfamiliar in the beginning and therefore difficult to answer. However, they reported feeling significantly more confident in completing the questionnaires after participating in the intervention.

> *“I felt a lot more equipped to answer those questions [at the end], and maybe because we went through all of the sessions, we kind of prepared for it as I’d thought about a lot of things throughout those sessions, so at the end it made it easier to fill them in.” (C2)*

#### Feasibility of assessing the cost of the programme

The total cost of delivering “Encompass” to eight participants was 4,400 Great British Pounds (GBP) (table 7), comprising 1,520 GBP in investment costs and 2,880 GBP in recurrent costs. Recurrent delivery costs were 360 GBP per participant.

**Table 7:** Programme provider costs in 2024 Great British Pounds (GBP) (N=8)

| Cost category | Cost | % of total cost | Cost per participant |
| --- | --- | --- | --- |
| **Investment costs** |  |  |  |
| Facilitator training (Labour/ Personnel)<sup>1</sup> | 1260 | 29.9% | 157.50 |
| Printing manuals and resources (Materials/ Supplies/ Equipment) | 60 | 1.4% | 7.50 |
| Mats, equipment and toys (Materials/ Supplies/ Equipment) | 200 | 4.7% | 25 |
| Total investment costs | **1520** | **36%** | **190** |
| **Recurrent costs** |  |  |  |
| Facilitator time (Labour/ Personnel)<sup>1</sup> | 1980 | 42.7% | 247.50 |
| Venue hire (Facilities)<sup>2</sup> | 700 | 16.6% | 87.50 |
| Refreshments (Materials/ Supplies/ Equipment) | 200 | 4.7% | 25 |
| Total recurrent costs | **2880** | **64%** | **360** |
| Total cost | **4400** |  | **550** |

1. Includes proportion of salary for one NHS healthcare professional aligning with NHS agenda for change pay scales (48) and one expert parent facilitator time aligning with NIHR payment guidance for members of the public involved in research (49). 2. Locally community library rental

Recording the costs of delivering “Encompass” was feasible but required careful tracking by the researcher. Data relating to costs were collated at the end of the study using the Childhood Cost Calculator, which provided a structured but relatively limited framework for capturing key categories such as staff time, materials and venue hire. To capture the costs of an intervention like “Encompass” at this early feasibility phase, a tailored spreadsheet-based approach would be sufficient.

The main challenge was determining exact costs for NHS staff time and navigating the contractual and funding arrangements between the university and NHS partners. This speaks more to the feasibility of conducting the research than to the feasibility of implementing “Encompass” itself as an intervention. Questions remain as to how to sustainably employ expert parents within the NHS. In a larger-scale evaluation, a potential cost-effectiveness framework would enable more systematic data collection and clearer cost attribution.

## Discussion

This study offers the first evidence that a participatory caregiver support programme originally developed in low- and middle-income countries can be feasibly delivered and evaluated within a diverse UK community health system setting. By testing “Encompass” in a real-world setting, we demonstrate that the adapted programme translates across vastly different contexts, and that caregivers and facilitators find it acceptable and relevant. This early proof of principle study demonstrates that key processes for the implementation and evaluation of the programme, such as facilitator training, fidelity monitoring and outcome data collection are feasible, with some amendments required for recruitment processes.

Key strengths of the study include meaningful involvement of parents with lived experience throughout, the inclusion of multiple stakeholder perspectives to support triangulation of findings at an individual, organisational and system level, and testing of core implementation processes (e.g. recruitment, training, fidelity) to inform feasibility and scalability within existing services.

The main limitation was the single-site design, which may limit transferability of feasibility findings to other settings or populations, even within high-income countries. The research team coordinated and closely oversaw delivery, which supported exploration of the feasibility of evaluating “Encompass”, but may limit understanding of how feasible implementation would be under routine service conditions.

### Comparison to other studies

Our findings broadly align with the UK-based proof-of-principle study of the “Healthy Parent Carers” (HPCs) programme (24), which reported similar strengths, limitations, and recruitment challenges. Despite differences in setting and population, both are group-based, lived-experience–facilitated programmes aiming to improve caregiver wellbeing. While recruitment was challenging in the initial HPCs study, a subsequent feasibility randomised controlled trial successfully achieved target numbers despite ongoing barriers (25).

The greatest challenges to diverse caregiver involvement in programmes like HPCs or “Encompass” are the practical constraints (e.g. time pressures, childcare, transport) and emotional or social barriers (e.g. feeling overwhelmed, guilt about self-care, feeling like the programme is not relevant to them) (50). Similar results were found in relation to recruitment and attendance challenges in group caregiver programmes in LMICs relating to time burden, competing obligations, stigma, social marginalisation, awareness gaps, and structural constraints (51–54). All the above-mentioned studies mention the importance of trusted messengers, community leaders or connectors, and local champions when addressing barriers to recruitment. Consistent with this, key partners emphasised the importance of understanding how different groups receive information, and caregivers highlighted peer word-of-mouth as a key strategy for improving recruitment and engagement

“Encompass”, “Baby Ubuntu” and “Juntos” all originate from the same participatory caregiver-support model, adapted for use in different countries and health systems. As programmes with common foundations but varied implementation contexts, comparisons provide useful insight into the feasibility and acceptability of delivering this type of group-based support. The group format and participatory materials were highly acceptable to “Encompass” participants, echoing findings from “Baby Ubuntu” (51) and “Juntos” (52,53).

Collecting data in our study relating to caregiver wellbeing and empowerment was feasible, although some caregivers required support to complete questionnaires. This again mirrors challenges reported in “Baby Ubuntu” and “Juntos”, highlighting the importance of simple, clearly explained measures when evaluating the impact of caregiver programmes in diverse communities. In particular, some of the “Encompass” caregiver participants found elements of the Family Empowerment Scale (FES) difficult to interpret, a difficulty also highlighted in a recent systematic review showing that some items on the FES may not be well-understood outside the original US context (55). In comparison, the Parent-Patient Activation Measure (P-PAM) was generally well-received across our diverse caregiver group, aligning with evidence from cross-cultural validation work showing that P-PAM has clear, accessible wording and good acceptability among parents in paediatric settings (56).

### Implications for Clinicians or Policymakers

The strong acceptability of “Encompass” appears to be driven by features that are consistently valued across similar caregiver programmes. Across “Baby Ubuntu” and “Juntos”, as well as programmes such as “E-PAtS” and “Healthy Parent Carers”, caregivers emphasise the importance of learning alongside others with lived experience, practical and relevant content, and group formats that feel inclusive and flexible. These studies also highlight common feasibility considerations, including practical barriers such as childcare, transport and competing appointments.

Questions remain as to whether “Encompass” is feasible to deliver to multiple groups of caregivers in different areas of the UK within an NHS context. A larger sample size and comparison group would allow for more meaningful evaluation of effectiveness on caregiver wellbeing and empowerment. Further exploration is also needed regarding online or hybrid delivery. Finally, although this study focused on early years, caregivers of children with complex neurodisability require support at multiple transition points. There remains a gap in provision for families at later stages, such as transition to secondary school and adult services.

## Conclusions

A co-designed group programme originating from low- and middle-income countries, that supports caregivers of children with complex neurodisability, can feasibly be delivered and evaluated in an ethnically and linguistically diverse UK community health setting. This study provides an initial proof of principle of the feasibility of delivering the programme, with all progression criteria being met or requiring only minor amendments. “Encompass” supports peer learning, shared problem-solving, and early family-centred intervention, with potential to improve outcomes for children and families.

## Supporting information

Encompass feasibility supplementary files

## Statements

### Data availability statement

The data that support the findings of this study are available on request from the corresponding author. The data are not publicly available due to privacy or ethical restrictions.

### Ethics approval statement

The study was conducted in accordance with the Declaration of Helsinki and approved by the NHS Health Research Authority ref. 23/EM/0213, approval date 4 January 2024.

### Patient consent statement

Informed consent was obtained from all participants involved in the study.

### Clinical trial registration

ClinicalTrials.gov Identifier: NCT06310681

## Acknowledgements

We would like to thank members of the Encompass advisory group for their ongoing support and expertise throughout the research project including Frances Badenhorst, Dr Aleksandra Borek, Dr Phill Harniess, Alea Jannath, Rachel Lassman, Prof Christopher Morris, Rachel Osbourne, Prof Cally J Tann, Assoc Prof Tracey Smythe, Keely Thomas, Melanie Whyte and Dr Emma Wilson. Thank you to the mothers who generously gave their time to participate in this study.

## Author contributions

KP, AH, and MH conceptualised the paper and were involved in funding acquisition. The methodology was developed by KP, KB, CH, CJT, MH and AH. Data were collected by KP and analysed by KP, KB, CJT, MH and AH. The project administration was undertaken by KP, with AH, MH, KB, and CH supervising. KP, KB, MH and AH drafted the original manuscript, and all authors contributed to reviewing and editing it.

