## Supplementary material for "Feasibility of an adapted participatory group programme for caregivers of children with complex neurodisability in the United Kingdom: Results from the Encompass-2 study": Encompass feasibility supplementary files

Supplementary file 1 – Fidelity checklist

**ENCOMPASS Fidelity Checklist**

Quantitative tick-box metrics were developed for various intervention components, which were based on a Fidelity Checklist developed and utilised by the Baby Ubuntu implementers and researchers globally.

Data will be collected via researcher observations for each group and four randomly selected modules will be attended by an external researcher who will complete the fidelity checklist.

Fidelity is defined as:

≥ 70% delivery on items described in the checklist below relating to logistics, preparation and facilitator skills.

| **Date of Group:**  **Location:** | | | | | **Facilitators:**  **Start time: End Time:** | | | | | |
| --- | --- | --- | --- | --- | --- | --- | --- | --- | --- | --- |
|  | **Activity demonstrated** | | | | **Yes** | **No** | **Comment** | | | |
| **Logistics** | 1. The group was held at the date, time and location that was expected | | | |  |  |  | | | |
|  | 2. Number of facilitators who attended was the same as expected | | | |  |  |  | | | |
|  | 3. Number of parents/carers who attended were the same as expected | | | |  |  |  | | | |
|  | 4. All equipment needed for the session was present | | | |  |  |  | | | |
| **Preparation** | 5. Room was set up to include all parents/carers and children in a circle | | | |  |  |  | | | |
|  | 6. Facilitators greeted group, introduced themselves, and introduced the topic for the session | | | |  |  |  | | | |
|  | 7. Facilitators allowed parents/carers to introduce themselves (this may be part of an ice breaker activity) | | | |  |  |  | | | |
|  | 8. Facilitators ask for the group’s participation to construct group guidelines (e.g. confidentiality, non-judgement, kindness, active listening) in the first session and to refer back to these at the start of each subsequent session | | | |  |  |  | | | |
| **Facilitation skills** | | **Facilitator expert parent:**  **Name:** | | | | | | **Facilitator health professional:**  **Name:** | | |
|  | | Y | N | Comments | | | | Y | N | Comments |
| 9. Facilitator followed the content of the manual, completing ice breaker and all components of the module | |  |  |  | | | |  |  |  |
| 10. Facilitator was prepared and knowledgeable of the content (i.e., speaking with ease, correct information, and not only reading directly). | |  |  |  | | | |  |  |  |
| 11. Facilitator used demonstration and equipment to communicate to parents and included parents in activities | |  |  |  | | | |  |  |  |
| 12. Facilitators asked parents/carers to share their own experiences of a specific topic, encouraging a participatory approach   (e.g. asking about parents/carers’ different experiences relating to feeding, or if a parent/carer asks a question, to sometimes ask it back to the group) | |  |  |  | | | |  |  |  |
| 13. Facilitator summarised discussions | |  |  |  | | | |  |  |  |
| 14. Facilitator asked the group if they had any questions after an activity and allowed time for discussion as needed | |  |  |  | | | |  |  |  |
| 15. Facilitator encouraged all members of the group to participate (without forcing) and managed quieter/ more dominant group members | |  |  |  | | | |  |  |  |
| 16. Facilitator used positive verbal and non-verbal communication and body language (e.g. nodding, leaning forwards, smiling) and thanking any group members for their participation | |  |  |  | | | |  |  |  |
| 17. Facilitators worked as a team and supported each other (e.g. referring to the other if they did not know how to answer a question, or supporting a group member if they become upset while the other continues) | |  |  |  | | | |  |  |  |
| 18. Facilitators ended the group by summarising what was spoken about and thanking the group members for their participation | |  |  |  | | | |  |  |  |

Supplementary file 2 – Topic Guides

**Caregiver participants**

Guide for interviewer:

- Bullet pointed items are key questions to ask – these are open questions wherever possible.
  - *They are followed by suggested probes to use to obtain more detail from the participant – the interviewer may not need to use these probes if the participant has already spontaneously discussed these particular aspects of their experience*
- The questions become more specific as each section progresses - the interviewer may not need to ask later bulleted questions in each section if the participant has already covered them earlier in the conversation

**General experience of the Encompass groups**

- What was your experience of attending the Encompass groups?
  - What did you like/dislike about attending the groups?
  - Were there any barriers to attending the groups?
- How did you find the different module topics?
  - Which ones were more relevant for you and which ones were less so?
- When and how did you first find out about the groups?
  - What made you decide to join?
  - Did you have any concerns or apprehensions before joining? How did those change/remain as the groups progressed?
  - Do you have any ideas about how to make the groups more inclusive? To reach a wider group of people?
- How did you find the facilitation?
  - What was your experience of having two facilitators with different skill sets and backgrounds?
  - What did the facilitators do that was helpful?
- How did you feel going to the groups?
  - What were your thoughts and feelings before you went each session? Did this change at the end of a session?
  - What motivated you to go?
  - How did you feel during the group sessions?
  - Were there any negative aspects to the groups?
  - Why do you think some parents/carers may not attend these groups?

**Outcome measures/ questionnaires**

- How did you find filling out the questionnaires at the beginning and end of the group programme?
  - Which ones were easier to fill out? Which ones were more difficult? If so, why?

**Capability**

*TDF – knowledge, skills, memory, attention and decision processes*

- Have you seen a change in yourself since attending the groups?
  - For example, how you understand things, or changes in routine etc.
- What were the most important things you learnt from the groups?
  - Are there any new skills that you have gained?
  - Can you provide examples of how the groups may have influenced your understanding of your child's disability and their specific needs?
  - Are there any gaps in your knowledge? How could the group have addressed these?
- How easy or difficult have you found carrying out what you learnt from the groups?

**Opportunity (environmental factors)**

*TDF – environmental context and resources, social influences*

- What helped or hindered you from attending the groups?
  - What barriers might other parents/carers have faced in attending the groups?
  - What could we do to address these barriers?
- How did you find sharing what your learned with your family and friends?
  - What was easier to share and what was more difficult?
  - How did they respond?
  - Do you feel that there are in changes as to how your family and friends view your child?
- Were there any barriers to carrying out the skills that you learned during the groups?
  - What made it easier or difficult to practise the skills that were learned in the group?
- How did you find connecting with other parents in the group?
  - Did you share similar experiences with other parents/carers?
  - Did you keep in contact between groups?
  - What helped you to connect? What made it difficult? (e.g. language)

**Motivation (attitudes and beliefs)**

*TDF – Beliefs about capabilities (confidence) and consequences, Intentions (I plan to), Goals (I want to), emotions*

- Have you noticed any changes in your confidence as a parent/carer after the groups?
  - If so, what might these be?
  - What might make you feel more confident?
- What do you think will happen if you continue to carry out the skills learnt during the groups?
  - What are your continued goals for your child?
  - What might the overall impact on your child be?
- How did the groups make you feel?
  - What parts of the groups might have helped or hindered wellbeing?
  - How do your feelings at the time affect whether you are able to carry out the skills learnt during the groups?

**Context**

*Context compass – ‘Fit’*

- When do you think parents/carers should be referred to the Encompass groups?
  - Straight after your child’s diagnosis? Or allow some time to pass?
- How do you think Encompass groups may work alongside / or with your current appointments – e.g. paediatrician /physio/OT/speech and language therapy?
  - What other professionals are you currently under?
  - Many parents/carers describe the burden / overwhelm of too many appointments – how might Encompass help or hinder with this?

**Significant Changes**

- How has your child been since the programme started?
  - Has anything changed? E.g. functional changes, child feeling happier, participating more in the community etc.
  - What has been the most important change for your child?
- Where do you go to get support if you have a problem with your child?
- What has been the most significant change for you as a parent/carer?

**Facilitator participants**

Guide for interviewer:

- Bullet pointed items are key questions to ask – these are open questions wherever possible.
  - *They are followed by suggested probes to use to obtain more detail from the participant – the interviewer may not need to use these probes if the participant has already spontaneously discussed these particular aspects of their experience*
- The questions become more specific as each section progresses - the interviewer may not need to ask later bulleted questions in each section if the participant has already covered them earlier in the conversation

**General Training and Delivery**

*TDF domain – knowledge and skills, beliefs about capabilities*

- How did you find the training for the Encompass groups?
  - What did you find useful?
  - What was not as useful?
  - What aspects of the training helped you to deliver the Encompass groups to the best of your ability?
  - Were there any gaps in the training that you would have liked?
- How confident did you feel in facilitating the groups?
  - What might have helped you to improve your confidence?
- How did you find delivering the Encompass groups?
  - How was the preparation for the groups each week?

What was easy and what was difficult?

- How did you find following the manual?
- How did you find the activities/ice breakers/demonstrations?
- How did you find the venue and equipment?
- How did you find being a facilitator?
  - How did you find having two facilitators?
  - How did you work together? What made it easier and what made it more difficult?
  - How did you find managing group dynamics?
- Overall, what could have been improved or changes about the groups to make them easier for you to deliver?
  - For example you said x was difficult – is there anything that could have been done differently to make it easier for you?

**Response of parents/carers**

*TDF – beliefs about consequences*

- Do you think the groups were helpful for parents/carers?
  - How relevant were they for parents/carers?
  - Did you notice any changes for parents/carers as the groups progressed?
- Was there anything difficult or unhelpful about the sessions for parents/carers?
  - Did you find that parents/carers showed signs of distress? If yes, did you do anything to help with that?
- Overall, what could have been improved or changed about the sessions to make them more helpful for parents/carers?
- Was there anything you would have liked the sessions to cover that wasn’t covered?

*TDF – environmental context and resources*

- Was there anything that got in the way of parents/carers attending the sessions?
  - What helped or hindered parents/carers to attend the sessions?
  - What could be done to reach more parents/carers? To make the groups more inclusive?
  - Were further resources required to make the groups run more effectively? What were these?

**Context**

*Context compass – ‘Fit’*

- What are the current priorities within community child health settings?
  - Does Encompass align with these priorities?
- When do you think parents/carers should be referred to the Encompass groups?
  - Straight after child’s diagnosis? Or allow some time to pass?
- How do you think Encompass groups may work alongside / or with current appointments – e.g. paediatrician /physio/OT/speech and language therapy?
  - Many parents/carers describe the burden / overwhelm of too many appointments – how might Encompass help or hinder with this?

*Context compass – ‘Readiness’*

- What would a community child heath setting need in order to run further Encompass groups?
  - E.g. Structural factors – e.g. physical space, computers, data, materials needed
  - E.g. Staffing – what about turnover/stress/overwhelm?
  - E.g. leadership – who would be able to lead a programme like this?
  - E.g. relationships – what team and inter department dynamics are needed?
- What are your views on whether it is feasible to use the ENCOMPASS groups in community child health services?

**Key partner participants**

Guide for interviewer:

- Bullet pointed items are key questions to ask – these are open questions wherever possible.
  - *They are followed by suggested probes to use to obtain more detail from the participant – the interviewer may not need to use these probes if the participant has already spontaneously discussed these particular aspects of their experience*
- The questions become more specific as each section progresses - the interviewer may not need to ask later bulleted questions in each section if the participant has already covered them earlier in the conversation

Begin by describing the Encompass group programme – a participatory community-based group programme for parents/carers of children with complex neurodisability under the age of 5 years. Facilitated by both a health professional and an expert parent/carer with lived experience. Emphasise the participatory nature of the group and that parents/carers learn from each other’s experiences. Briefly describe examples of modules.

**Context Compass**

*Fit*

- Could you describe how decisions are made in community child health services in this setting?
  - Is there an openness to change?
  - Is it flexible/ more rigid?
- What are the current priorities within community child health settings?
  - How do you decide what the priorities are?
  - Does Encompass align with any of these priorities?
- Is there a current tension for change?
  - Is the organisation looking to change the way they support children with complex neurodisability? Or are things going well?

*Readiness*

- What resources and funding are available within the NHS setting for new innovations?
  - If you were to introduce a new programme, would this involve re-distribution of costs or could funding be applied for?
- What partnerships and connections exist with the NHS community child health settings?
  - E.g. schools, social care, charities
  - Would there be incentive to link up with these partnerships? How might this look?
- What does leadership look like in the setting?
  - What capacity would leaders have to support implementation of Encompass?
- What are the relationships like within teams and between departments at this setting?
  - How have previous innovative projects worked?
- What does staffing look like?
  - Is there high turnover?
  - How are the waiting lists?
  - Is there staff capacity for a new programme such as Encompass?
- Are there support structures available within the setting who might assist in carrying out the programme?
  - E.g. QI
- What policies currently drive clinical practice in this setting?
  - How does Encompass align with these policies? Or not?

*Implementation/sustainability*

- What factors that we have discussed during this interview (e.g. partnerships, resources, supports) are the most critical if Encompass were to be implemented?
- What are the greatest barriers that you envision for ongoing implementation/ scaling up of Encompass?
- What might be required for ongoing monitoring of Encompass if it was implemented?
  - E.g. staffing, resources, continued support from researchers

*Characteristics of the Encompass programme*

- What would make community child health settings more likely to adopt Encompass?
  - Research evidence?
  - Adaptability of Encompass? To fit local needs
  - Design – well designed and packaged and presented?
  - Costs – associated with adopting Encompass into the setting
